# Well-being and self-care strategies for cardiovascular genetic counselors: a qualitative study

**DOI:** 10.1101/2025.04.10.25325617

**Authors:** Laura Yeates, Lucas A Mitchell, Ivan Macciocca, Helen Mountain, Mary-Anne Young, Colleen Caleshu, Alison McEwen, Jodie Ingles

## Abstract

**Purpose:** To explore the impact of cardiovascular genetic counseling practice on genetic counselor well-being and describe self-care practices.

**Methods:** Participants were recruited through the Australasian Society of Genetic Counselors. Semi-structured interviews explored challenges in cardiovascular genetic counseling practice, supervision and self-care. Interview transcripts were analysed using reflexive thematic analysis. Self-reported demographics, psychological well-being and burnout measures were used.

**Results:** Eighteen genetic counselors participated. Median interview length was 54 minutes (range 40-74). All participants were female and 83% of European ethnicity. Few reported mild or moderate depression symptoms (17%), mild or moderate anxiety symptoms (22%) and none (0%) had scores indicating stress. Three (17%) had scores indicating burnout. Thematic analysis identified three themes: (1) cardiovascular genetic counseling is different, not harder or easier; (2) workplace pressures affect well-being; (3) a self-care “tool kit” is necessary and supervision is a key component.

**Conclusion:** Genetic counseling practice and workload can affect well-being. A genetic counselor self-care ‘tool kit’ that includes supervision helps maintain well-being.

**Graphical abstract:** 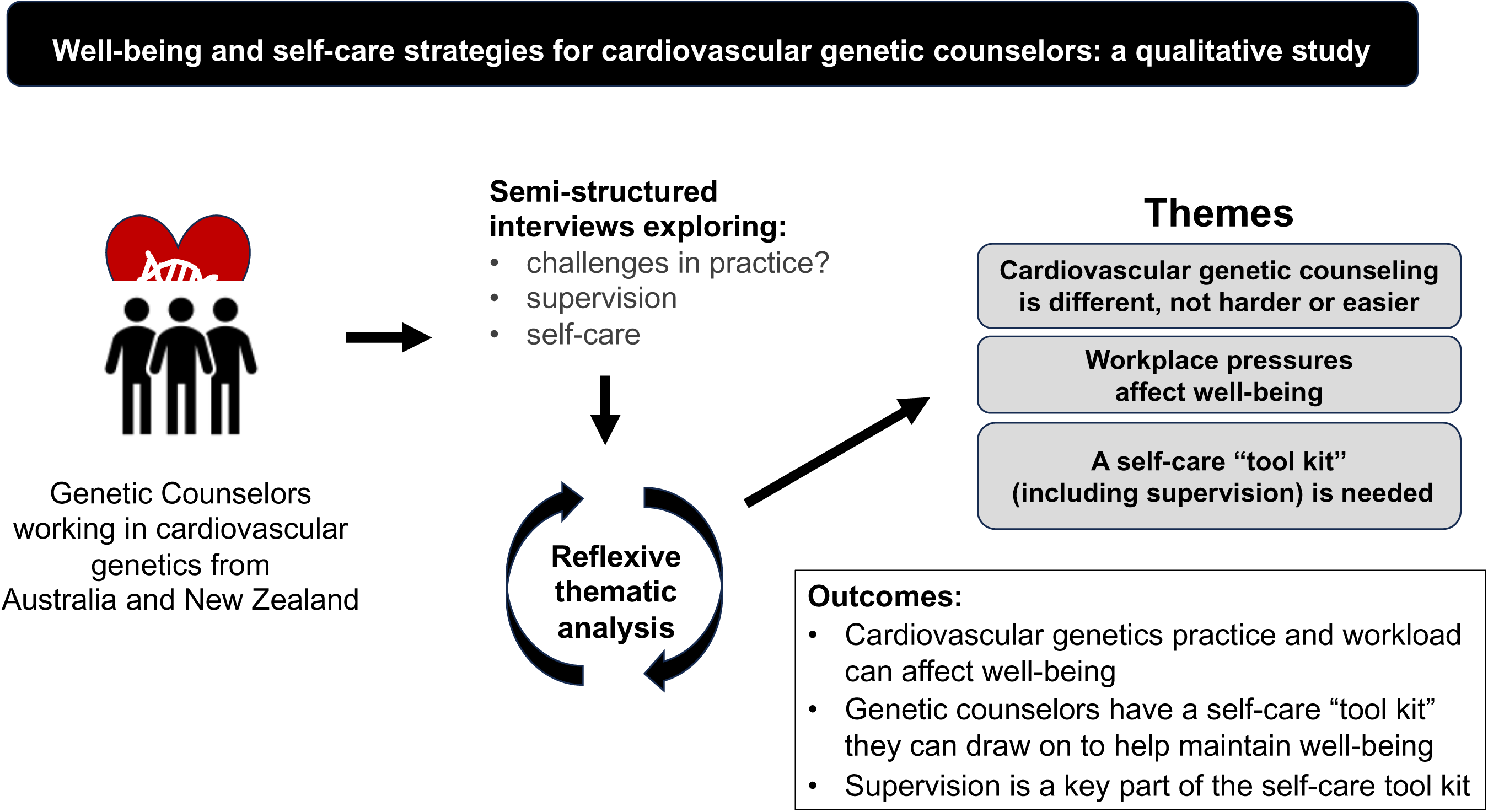

## INTRODUCTION

Working in “helping professions” like genetic counseling can impact healthcare professionals’ well-being due to repeated exposure to difficult clinical scenarios.^1^ Genetic counselors are at risk of burnout and compassion fatigue, which can occur after repeated empathetic engagement with those who are suffering.^2,3,4^ Burnout refers to exhaustion that comes from excessive demands on an individual.^5^ Previous studies report high levels of genetic counselor burnout and compassion fatigue.^2,6,7^ In a study examining occupational stress and burnout in North American and Australian genetic counselors, 41% had high levels of burnout.^5^ Likewise, a study of American genetic counselors by Caleshu et al reported 57% of participants had scores indicating burnout, with lack of administrative support associated with higher levels. Lower levels of burnout were associated with professional self-care behaviours.^3^ The American National Academy of Medicine developed a systems model of professional burnout, which highlighted job pressures including lack of administration support, workload and time pressures, and inadequate technology as key contributors to burnout.^8^

Clinical supervision is a learning process involving case discussion and reflection used in many healthcare professions, particularly in the fields of psychology and social work.^9–11^ Worldwide, genetic counseling regulation bodies include supervision as a requirement during training for genetic counselors, but following certification, requirements vary. In Australasia, career-long supervision is a requirement of practice for all genetic counselors.^12^ For Australasian genetic counselors, clinical certification is obtained after a minimum of one year fulltime employment (or equivalent) and completion of a certification portfolio.^12^ Supervision is mandated for genetic counselors who are yet to certify, including a minimum of 10 hours per year of individual supervision with additional group, peer supervision and case review up to 40 hours per annum. Once certified, genetic counselors must participate in a minimum of 10 hours per year of supervision (individual, peer or group) to maintain their registration.^12^ Inherited cardiovascular conditions are a subspeciality of genetic disease, involving care of families with inherited cardiomyopathies, arrhythmias, aortopathies, hyperlipidaemias and congenital heart disease.^13^ Due to clinical heterogeneity, patients can be asymptomatic, or experience syncope, heart failure and sudden cardiac death (SCD).^13^ Adjustment to a new diagnosis can be challenging, with the need to consider risk to family members, medication adherence, and in some cases an implantable cardioverter defibrillator, as well as sport or employment restrictions.^14,15^ Managing the risk of SCD can be especially difficult, leaving families dealing with uncertainty about ongoing risk to family members.^15,16^

Genetic counselors play a key role in assisting individuals and families to navigate the clinical, familial and psychosocial impacts of inherited cardiovascular conditions.^14^ Repeated exposure to difficult clinical situations has potential to impact genetic counselor well-being and contribute to burnout. We aimed to explore the impact of cardiovascular genetic counseling practice on well-being, the benefits and barriers of clinical supervision in maintaining well-being, and self-care practices of genetic counselors working in cardiovascular genetics.

## METHODS

### Participants

The study was approved by Human Research Ethics Committee of the University of Sydney [2023/003]. Participants provided written, informed consent. Participants were recruited from the Australasian Society of Genetic Counselors (ASGC) weekly e-newsletter. An advertisement ran for six weeks inviting ASGC members with current or previous experience in cardiovascular genetics to participate in a study exploring the impact of cardiovascular genetic counseling practice on genetic counselor well-being. Participants completed a questionnaire via REDCap^17^ including demographic details, type of inherited cardiac conditions seen, proportion of their clinical load taken up by cardiovascular genetic cases and frequency of clinical supervision. Participants identified their ethnic background from one of the nine categories derived by the Australian Bureau of Statistics.^18^ The questionnaire included the Professional Fulfilment Index (PFI), a validated scale that measures burnout and professional fulfillment;^19^and the Depression Anxiety Stress Scale short form (DASS-21), a validated 21 item questionnaire measuring depression, anxiety, and stress.^20^ DASS-21 depression scores assess concepts like ‘dysphoria’ and ‘hopelessness’, the anxiety scale assesses physical symptoms of anxiety and the stress scale assesses issues such as ‘chronic non-specific arousal’, ‘inability to relax’, ‘irritation’ etc.^20^ Information power was used to determine sample size, this practice considers five items: study aim, sample specificity, use of established theory, quality of dialogue and analysis strategy.^21^ Given our study aim was relatively narrow, our sample dense, analysis used an established theory, we had strong quality of dialogue and the analysis strategy was cross-case, this model led us to determine a minimum sample of 15 participants.^21^

### Study design

A semi-structured interview schedule was developed by the research team. Questions about the participants career, perceived challenges of working in cardiac genetics, workplace conflicts, engagement in supervision and self-care practices were included. Interviews were conducted via Zoom by LM, a research genetic counselor who had not worked in cardiac genetics and was least likely to have a pre-existing relationship with participants. Interviews were recorded and transcribed verbatim.

### Data analysis

DASS-21 scores were calculated to measure symptoms of depression, anxiety and stress and classified into one of five categories (normal, mild, moderate, severe, extremely severe).^20^ Professional Fulfilment Index scores were calculated for each of the three subscales (professional fulfilment, exhaustion and interpersonal disengagement). The exhaustion and interpersonal disengagement subscales were then used to calculate the burnout score. A cut-off of 14 or above was used to indicate burnout.^19^ Interview transcripts were de-identified and coded in NVivo. We used a grounded theory approach with iterative review of the transcripts using Braun and Clarke’s six stages of reflexive thematic analysis.^22^ LY and LM reviewed the first six transcripts independently and discussed preliminary codes. The remaining transcripts were coded iteratively by LY, with codes being continuously refined. Initial themes were developed and reviewed by JI, LM, HM, IM, AM and LY. Themes were then further defined and refined.^22^

## RESULTS

### Demographic information

Eighteen individuals consented to participate (Table 1). All were female with 10 (55%) being aged under 40 years. Fifteen (83%) identified as European and 10 (56%) had been working less than 10 years as a genetic counselor. Six (33%) participants had a cardiac case load over 50% with four (22%) who exclusively saw patients with cardiovascular disease. All participants (100%) had experience seeing patients with cardiomyopathy, 94% with inherited arrhythmia syndromes, and 83% with families after SCD of a relative. Participants were asked about current supervision attendance. All were participating in at least one supervision practice (Table 1).

**Table 1.** Demographics of the participants.

| **Demographics** | **n (%)** |
| --- | --- |
| **Sex** |  |
| Female | 18 (100) |
| Median interview length in minutes (range) | 50 (40-74) |
| **Age** |  |
| <39 years | 10 (55) |
| 40+ years | 8 (45) |
| **Ethnicity** |  |
| European | 15 (83) |
| **Family make up** |  |
| Partnered | 14 (78) |
| Children | 10 (55) |
| **Years working as a genetic counselor** |  |
| Less than 5 years | 5 (28) |
| 5 – 9 years | 5 (28) |
| 10 – 19 years | 5 (28) |
| 20+ years | 3 (16) |
| Additional training in grief and loss | 4 (22) |
| **Types of cardiac disease seen** |  |
| Cardiomyopathies | 18 (100) |
| Inherited arrhythmias | 17 (94) |
| Sudden cardiac death | 15 (83) |
| Aortopathies | 14 (78) |
| Familial hypercholesterolaemia | 6 (33) |
| **Supervision type – (any frequency)** |  |
| Individual | 13 (72) |
| Peer supervision | 12 (67) |
| Group supervision | 15 (83) |
| Provide individual supervision | 8 (45) |
| Facilitator of group supervision | 1 (6) |
| **DASS-21 (mild or moderate)** |  |
| Depression | 3 (17) |
| Anxiety | 4 (22) |
| Stress | 0 (0) |
| **Professional Fulfilment Index** | **Median (range)** |
| Fulfilment | 70 (45.8-100) |
| Exhaustion scale | 37.5 (0-56.25) |
| Interpersonal disengagement | 8.3 (0-37.5) |
| **Burnout** | **n (%)** |
| Proportion Yes (cut off 1.3) | 3 (17) |

Abbreviations: DASS-21 = Depression anxiety stress scale short form.

DASS-21 scores for depression, anxiety and stress were calculated and categorised into normal, mild, moderate, severe, and extremely severe.^20^ In total, 17% reported mild or moderate depression, 22% reported mild or moderate anxiety and none (0%) had scores indicating stress. To better understand these scores, including the seemingly low rate of stress (range 0-12), the highest rated items were determined. For the depression sub-scale, the item “*I found it difficult to work up the initiative to do things*” was most endorsed (n=11, 61% selected often/sometimes). For responses to the anxiety items, “*I felt that I was using a lot of nervous energy*” was rated the highest (n=7, 39% selected often/sometimes); and for the stress subscale the highest rated item was, “*I find it hard to wind down*” (n=14, 78%, selected almost always/often/sometimes). Three (17%) of the cohort had professional fulfilment scores indicating burnout (Table 1).

### Thematic analysis

Thematic analysis of the transcripts identified three themes: 1) cardiovascular genetic counseling is different, not harder, or easier; 2) workplace pressures affect well-being; 3) A self-care tool kit is necessary, with supervision a key component (Figure 1).

**Figure 1-.**
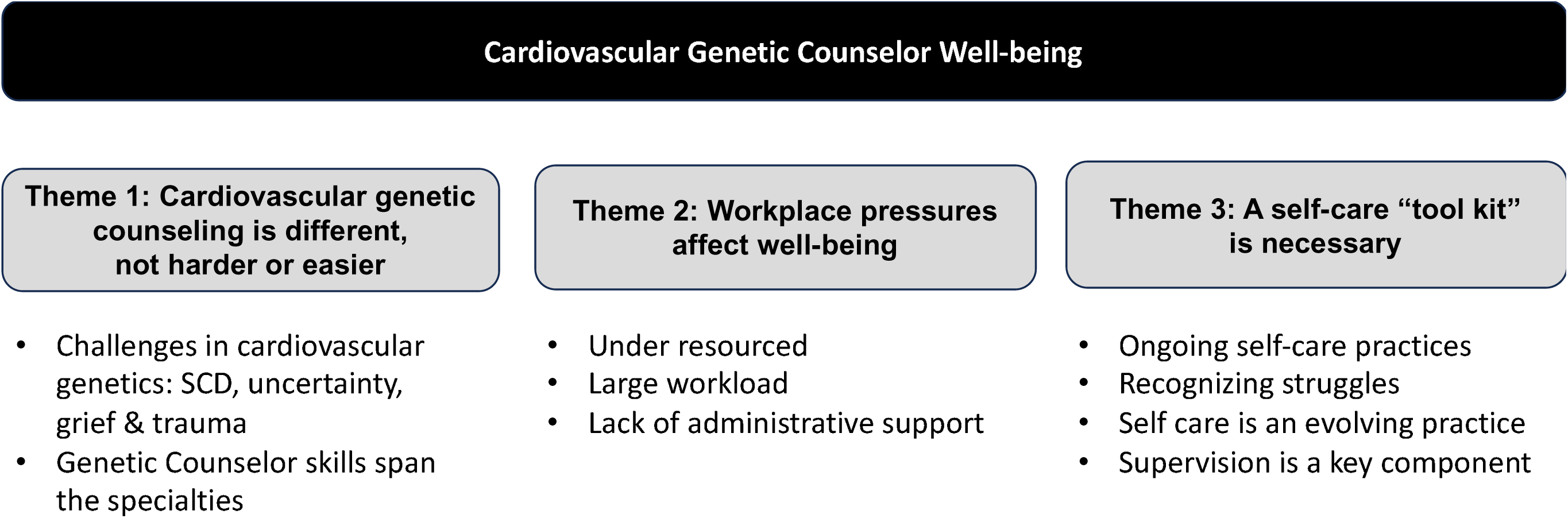
overview of themes. abbreviations SCD = sudden cardiac death.

**Theme 1: Cardiovascular genetic counseling is different, not easier or harder.**

Participants described the benefits of working in cardiac genetics, as a relatively new area of practice where they could see the direct advantages of testing and identifying at-risk individuals.

> *“I think it’s really rewarding where you can be that support person, but also the testing can make difference in the management of patients and save lives.”* Participant 3, <50% cardiovascular genetics caseload

Participants liked being on the forefront of a relatively new area in genetics and appreciated the opportunity to work within a multidisciplinary team and contribute to variant curation.

> *“I definitely say it’s fascinating and like it’s a really wonderful part of genetic counseling to get to be involved in the genetics itself is really interesting…it’s a good opportunity to learn about variant curation”* Participant 10 <50% cardiovascular genetics caseload

Participants described the challenges associated with cardiovascular genetics practice (Table 2). In part, these challenges related to dealing with uncertainty, which included lack of penetrance data about pathogenic variants, the types of clinical presentations and outcomes patients might expect throughout their lifetime, including the inability to predict who might suffer from heart failure or SCD. Participants described the difficulties working with families after SCD. Given the sudden onset of these events, often without warning, surviving family members often experience trauma and grief requiring support from an appropriately trained person.

**Table 2.** Challenges of cardiovascular genetics.

| **Challenges of cardiac genetics** | **Examples** |
| --- | --- |
| **Clinical impact of disease** | Adjusting to new diagnosis |
|  | Sports restriction |
|  | Uncertainty in clinical outcome |
|  | Clinical heterogeneity |
|  | Risk of sudden death/ fear of event |
|  | Grief and Loss |
|  | Trauma |
|  | Lack of answers |
| **Continuing education** | Upskilling in cardiac genetics |
|  | Variant curation |
|  | Grief and loss |

“*The uncertainty is an important aspect to the genetic counseling. The fact that we can’t predict the outcome or how people are going to progress with their disease, the fact that there’s no “cure”, nothing you can do, unlike in cancer, to prevent something from happening, you can just manage it…if it comes about”* Participant 17, ≥50% cardiovascular genetics caseload

> *“you know, when you’re dealing with someone whose child goes out for a walk and then they don’t come home, um, that’s hard.”* Participant 6, <50% cardiovascular genetics caseload

The lack of warning signs with SCD can affect the genetic counselor, sometimes causing countertransference or thoughts about their own family.

> *“Even though I understand the statistics and how rare these events are, I personally find it difficult… most of the time I’m okay but removing myself and my family from the situation and having I guess a confidence that it’s not going to happen to us can be difficult when you’re seeing it every week.”* Participant 17, ≥ 50% cardiovascular genetics caseload

While participants recognised some challenges were unique to cardiovascular genetics, such as caring for families after SCD, they highlighted that there are difficult aspects to all areas of genetics. The skills of genetic counsellors in dealing with uncertainty are necessary for many areas of genetics.

> *“Cardiac stuff is bad, don’t get me wrong, like sudden death awful. That sort of looming over your head. That’s a long life to live with that anxiety. But we also see a lot of other shit.”* Participant 14, <50% cardiovascular genetics caseload

> *“Similar to a prenatal setting when you’re dealing with…a structural anomaly found on scan for a foetus… these families want answers, and they need them quickly… and they’re pretty stricken and worried and there’s trauma there.”* Participant 13, <50% cardiovascular genetics caseload

> *“So it’s [prenatal] very different type of counseling compared to cardiac. But I mean it’s, it’s the same skill load I’d say”* Participant 9, >50% cardiovascular genetics caseload

**Theme 2: Workplace pressures affect well-being**

Regardless of speciality, the general workload impacted genetic counselor well-being. Participants identified their workload as a source of stress, which is sometimes compounded by on-call duties, availability of other members of the multi-disciplinary team (who may be part time) and the pressure of a long waiting list.

> *“We’re pretty understaffed…so I think my caseload is constantly too much… but I’ve just got to sit with the fact that people are waiting and they’re waiting on me to action things, but I’m only one person and I only have a certain amount of time in a day. So… it is hard and I always worry about people that are on the list don’t know their risk”* Participant 7 ≥ 50% cardiovascular genetics caseload

> *“there are times where the workload is very overwhelming. I think there was a time where our service was quite under-resourced and the counselors that were on the ground were very stretched… there were times where we would get together and cry because of the workload”* Participant 15 <50% cardiovascular genetics caseload

In addition, lack of administrative support impacts workload and in turn genetic counselor well-being. Participants spoke of having to help with administrative workload when other staff were unavailable, impacting the time available to complete their genetic counseling work and contributing to feelings of being overwhelmed.

> *“If there’s not enough administration support, the administration tasks end up on the genetic counselors, which gives us less time to focus on the counseling side of things. And…something kind of has to give… in some way. And either that is the counseling or the admin tasks fall to the wayside, neither is good for the patients or their care or their family’s care. And that kind of constant feeling of overworked is not good for the counselors or the team in general.”* Participant 5, <50% cardiovascular genetics caseload

**Theme 3: A self-care tool kit is necessary**

In considering the difficult clinical situations facing families, participants recognised that to some extent they “carry the cases with you” which led to feelings of being overwhelmed.

> *“There’s always one or, you know…there’s always a case that for some reason or another…might just sit with you in a different way for a bit longer.”* Participant 11, <50% cardiovascular genetics caseload

> *“most of the time I can compartmentalize my job. Um, but there have been moments where I haven’t been able to… sometimes you get these cases where someone’s lost a child, you just take that grief home with you, and it can be quite overwhelming carrying that at times.”* Participant 2, <50% cardiovascular genetics caseload

Participants spoke about a variety of self-care practices that they utilised throughout their career, which can be described as a self-care “tool kit” (Table 3). This included informal activities such as exercise, hobbies, seeing friends and family, ensuring adequate sleep, and formal activities such as supervision. The list of self-care practices reflected activities that participants dip in and out of as required, noting they are particularly important in stressful times. There were also certain practices they try to avoid like excess alcohol or “comfort foods” and working overtime.

**Table 3.** The self-care “tool kit”.

|  |
| --- |
| **Informal self-care: add to these and rotate as needed** |
| • Activities to decompress: cooking, hobbies, spending time with pets<br>• Boundaries between work and home (using commute to decompress)<br>• Eating well and meal preparation<br>• Exercise (including yoga)<br>• Informal debrief with colleagues<br>• Leaving work on time<br>• Recognizing limits/ saying no to extra activities that take time (at work or home)<br>• Recognizing life circumstances (e.g. children) can change what self-care looks like<br>• Restricting alcohol<br>• Routine is helpful in maintain self-care practice<br>• Seeing friends/ social life<br>• Self-talk – recognising when circumstances are out of your control<br>• Sleep |
| **Formal self-care** |
| • Annual leave/ holidays<br>• Professional counseling<br>• Supervision |

> *“It’s easy to sit there and kind of use food as a comfort zone and it’s maybe not the healthiest in the long run… like alcohol obviously is another thing that often I think people can turn to as a comfort and just a kind of ease of throwing the day away”* Participant 5, <50% cardiovascular genetics caseload

> *“I think when your workloads, when you know that you’re going through a stressful period in work, that’s when you have to actively pay attention to those other things. Whether it’s, you know, your diet, your social life, your family.”* Participant 9, ≥50% cardiovascular genetics caseload

Boundaries between work and home were important. Participants described activities to help them maintain boundaries, e.g., commute time, or activities to help decompress.

“*And then a nice long commute home where I often find myself reading or listening to a podcast, which just takes you out of your own head, makes you think about something else.”* Participant 4, ≥ 50% cardiovascular genetics caseload

> *“I think particularly for me, what has changed over time has been that ability to better separate work in my personal life…, it was very blurry in the beginning years and I would constantly think about work… nowadays what’s really helped me is…I keep work at work and I come home and I don’t think about work for the most part.”* Participant 13, <50% cardiovascular genetics caseload

In addition, participants described activities they may undertake when they notice they are not coping with their workload. Additional activities included the “informal debrief” with a colleague, and taking a break or annual leave. Participants spoke about informal debriefing providing a bridge to their next clinical supervision session, which may be a few weeks away. An “open door policy” in their workplaces enabled “ad hoc” informal debriefing.Participants felt they could utilise this as needed and appreciated the team approach to provide a listening ear.

> *“The debriefing, it’s, uh, we’ve got pretty much an open-door type scenario, so if you need to chat to someone, you can generally find someone to talk to.”* Participant 3, <50% cardiovascular genetics caseload

> *“When something hits… and like I’m not parking that [putting it on hold] until I see this person [supervisor] in two weeks’ time, I’m not doing it…because it needs to be addressed right now.”* Participant 6, <50% cardiovascular genetics caseload

### Supervision is a key component of the self-care “tool kit”

Individual, group or peer supervision was recognised as an important part of maintaining well-being. Supervision provided an opportunity for debriefing about difficult cases, and a space for self-reflection and learning.

> *“I think supervision is a really important part of being a genetic counselor…to learn from others, to be able to debrief in a safe space… to feel supported and, you know, to have that trust in your colleagues.”* Participant 12, <50% cardiovascular genetics caseload

Challenges among all types of supervision formats were discussed. Firstly, it was noted that there were difficulties in finding an individual supervisor with appropriate training (to meet board requirements) but also someone they would feel comfortable with. Feeling comfortable within group supervision was important. Participants recognised the vulnerability that comes with disclosure during supervision sessions and that building a supportive supervisory relationship was critical in maximising the benefits of supervision.

> *“I feel like <laugh> with group supervision, I haven’t really built that rapport yet and I feel um, a bit hesitant, like one occupying the space even though it’s meant to be there for that reason. But also too, just being able to comfortably talk about these things, I think I get a bit intimidated as well they’re like people who have been working for ages.”* Participant 16, ≥ 50% cardiovascular genetics caseload

*It’s definitely a nice space that everyone’s very supportive and you know, I don’t have any qualms, there’s no kind of conflicts that I’ve noticed arise…It’s just, I don’t feel fully comfortable being vulnerable in front of them.”* Participant 4, ≥ 50% cardiovascular genetics caseload

An additional barrier to supervision was the time taken to attend sessions, with participants identifying workload as a major barrier to attending supervision. For many, they put the demands of the clinic over their own self-care.

> *“Definitely when things have been really stressful with like caseload, there’s the odd time you’re just like, I don’t have an hour. Like this [attending supervision] just means that I’m going to have to work an hour later tonight… And it feels like an hour in the day that I don’t have”* Participant 10, <50% cardiovascular genetics caseload

Based on these responses, we created a roadmap to help both genetic counselors and their supervisors, to reflect on current self-care practices (figure 2) and promote an open dialogue about the impact of genetic counseling practice on well-being.

**Figure 2-.**
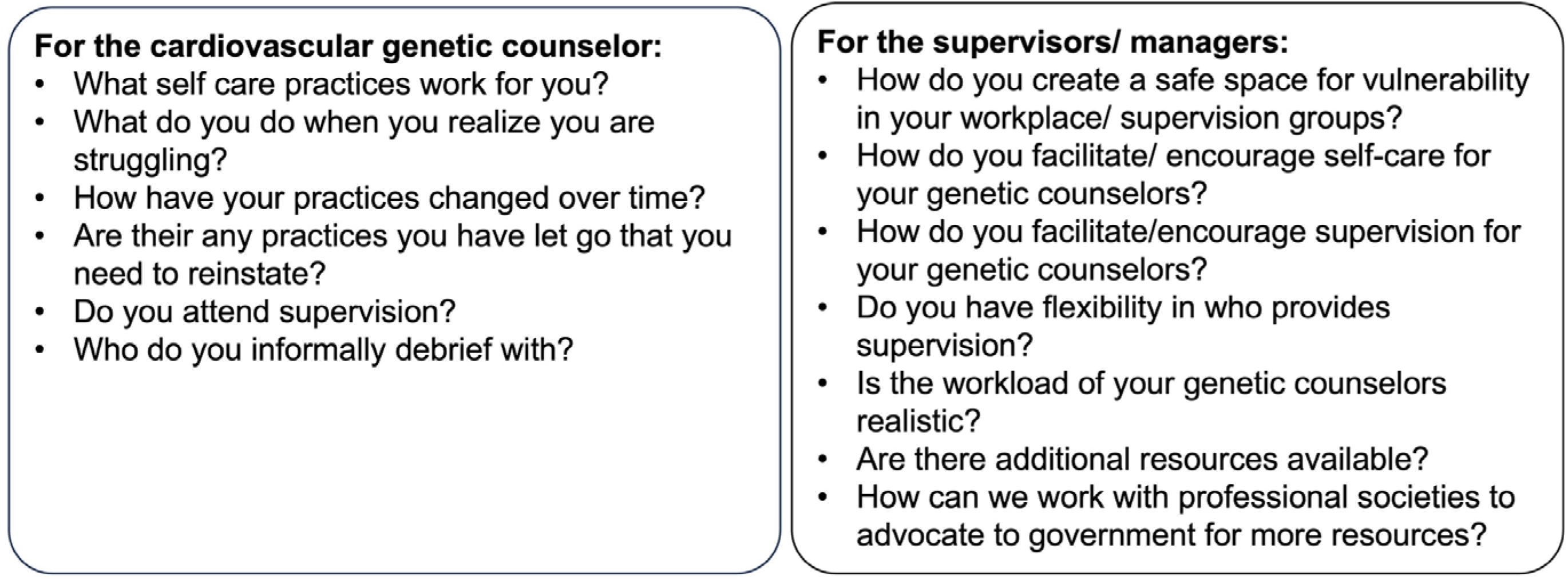
self-care road map for cardiovascular genetic counselors and supervisors

## DISCUSSION

Genetic counselors manage families with complex experiences and needs, exposing them to repeated empathetic engagement that can affect their well-being.^4^ In cardiovascular genetics, this includes managing families who have experienced the SCD of a young relative. We explored the impact of cardiovascular genetic counseling practice on genetic counselor well-being. Although aspects of the patient experience for those attending cardiovascular genetic clinics can be profoundly challenging, genetic counselors considered these to be no more or less challenging than in other clinical genetic settings. Genetic counselors’ training and skills equipped them regardless of the disease area they were practising in. High workload was frequently identified as negatively impacting on genetic counselor well-being, but engagement in self-care practices, including supervision, helped maintain well-being.

Among the genetic counselors who participated, there were relatively low levels of burnout reported, with 17% (3/18) having PFI scores indicating burnout. In contrast, a USA based study, found that 57% of responding genetic counselors had PFI scores indicating burnout.^3^ Participants in the USA study were recruited to a randomised controlled trial to evaluate professional well-being with and without mindfulness activities, potentially leading to selection bias whereby those genetic counselors struggling with well-being were more likely to engage with the study.^3^ Johnstone et al measured burnout in a sample of genetic counselors from USA, Canada and Australasia. Using the Maslach Burnout Inventory general survey, 47% of genetic counselors had moderate to high range burnout scores, with burnout being associated with occupational stress. While predictors of burnout were assessed, country of origin was not identified.^5^ Although the reasons for the differences in prevalence of burnout between our work and others are outside the scope of this study, considerations to the difference in scores is important noting the small sample size of our current cohort. There is the possibility of a biased sample, whereby those with low stress and burnout were more willing to participate in this current study. However, the lower levels of stress and burnout in this study group may also be due to self-care behaviours and the positive effect of mandatory supervision required for clinical practice in Australasia. Further work in a larger sample is needed to determine the true incidence of burnout in Australasian genetic counsellors and explore contributory and protective factors both in Australasia and internationally.

The importance of self-care was highlighted by our participants. Self-care has been defined as a range of activities performed by a person to promote well-being throughout their life.^23^ Self-care practices vary between individuals due to personal preference, life stage and resources available.^24^ Tension between the time needed for self-care practices, including supervision, and fulfilling clinical duties exists, and was highlighted by our participants. This included time for self-care practices outside the workplace such as exercise and hobbies. At times of stress or feeling an inability to cope, our participants reflected on their current self-care practices and increased the frequency, reinstated activities that had lapsed or sought additional support, either with an informal debrief with a colleague or in supervision. Although time away from clinical duties is needed to attend supervision, regular attendance and active participation in supervision enhances well-being and contributes to workers remaining in their field.^25^ Indeed, previous studies highlight the utility and importance of supervision in training of genetic counselors.^26–28^ For disciplines such as psychology and social work, the ongoing practice of supervision throughout a career provides an avenue to help maintain well-being and has been shown to promote retention.^10,25^

The importance of the supervisor/ supervisee relationship, or relationship between group members, were highlighted as a challenge by our cohort. Case reflection requires a level of vulnerability, opening oneself up to critique from others.^9^ For some participants, supervision was a supportive experience, while others reported barriers to vulnerability and disclosure. In the setting of genetic counseling supervision, it has been proposed that regular assessment and reflection of supervision practice may aid group dynamics.^29^

We found high workloads and lack of administrative support are detrimental to genetic counselor well-being, supporting Caleshu et al., finding that lack of administrative support was a predictor of burnout among genetic counselors.^3^ Our participants noted that when administrative support was lacking, they felt the pressure to step in and assist, taking them away from their core genetic counseling practice. Whilst budget constraints limit administrative support, the long-term consequences of failing to address this issue could be debilitating for the profession. In a profession experiencing workforce shortages both locally and internationally, where demand is predicted to grow,^30–32^ it is critical that employers address the lack of appropriate staffing and administrative support to enable genetic counselors to perform at their peak of practice. By providing genetic counselors with adequate administrative support, reasonable workloads and protected time to participate in well-being behaviours such as clinical supervision, we can ensure retention of a highly skilled genetic counseling workforce.

Consideration of digital tools and genetic counseling assistants^33,34^ that promote efficient service delivery and support core genetic counseling practice are warranted. A recent study assessed a webinar to provide pre-test counseling in hypertrophic cardiomyopathy and showed this to be an acceptable replacement for genetic counselor appointments, saving an average of 24 minutes, per patient.^35^ Other studies have used ‘chatbots’ for pre-test genetic counseling^36^ or facilitating communication to at-risk family members.^37^ Providing accurate online information that is accessible to patients with varying health literacy levels is important^38^ and has the potential to reduce face-to-face consultation time. Innovative service delivery and use of emerging technologies has the potential to create time for more complex discussions.

### Limitations

There were a number of limitations of this study worth noting. Firstly, this was a self-selected group, where cardiovascular genetic counselors experiencing burnout or poor well-being may have declined to participate. Secondly, our study invitation sought participants with any previous or current experience in cardiovascular genetic counseling, yet only the amount of current cardiovascular load was recorded. Thirdly, no participants had a DASS-21 stress score above the normal range. This could be due to small sample size or may be a reflection of the way stress is measured by DASS-21, as it would be expected that genetic counselors experience some stress. Finally, the cohort was homogenous, being all female and largely from a European background, and is reflective of the membership base of the ASGC where the participants were recruited from.^39^

### Considerations for future practice

Well-being is an important factor for ongoing genetic counselor practice regardless of speciality. To retain the workforce and ensure a long and fruitful career, genetic counselors must consider their self-care, recognising these practices can change over time and with life circumstances. Our study highlights key self-care practice considerations for cardiovascular genetic counselors. In addition, it identifies opportunities for supervisors and managers to reflect if they are providing adequate resourcing to their staff, with realistic workloads, additional administrative support, and facilitating access to supervision. Finally, further work to address funding issues is critical, with a need to work with professional societies to advocate to government for adequate funding and resources to provide services to meet demand (Figure 2).

## CONCLUSION

Well-being is an important consideration for all genetic counselors including those working in cardiovascular genetics. Genetic counsellors work in a range of specialties, and practice can impact the emotional well-being of genetic counselors. Workplace pressures and under resourced services place additional stress on genetic counselors. Ongoing self-care practice includes a range of activities that genetic counselors can individualise and build on over time with supervision being a key part of ongoing self-care.

## Data availability statement

The interview schedule can be requested from the corresponding author. Anonymized redacted transcripts can be made available upon request and with appropriate agreements and human research ethics committee approval.

## Author contributions

Conceptualization: L.Y., J.I., A.M., I.M., H.M., C.C. Methodology: L.Y., J.I., A.M., I.M., H.M., C.C. Recruitment: L.Y. Acquisition of the data: L.M. Data curation: L.Y., L.M. Formal analysis: L.Y., L.M., I.M., H.M., A.M. Drafting of the manuscript: L.Y. Critical review of the manuscript: L.Y., L.M., I.M., H.M., M.A.Y, C.C., A.M., J.I. Final approval: L.Y., A.M., J.I.

## Ethics declarations

This study was approved by the Human Research Ethics Committee of the University of Sydney [2023/003]. Participants provided written, informed consent.

